# Early adulthood socioeconomic trajectories contribute to inequalities in adult diet quality, independent of childhood and adulthood socioeconomic position

**DOI:** 10.1101/2024.05.07.24306992

**Authors:** Yinhua Tao, Jane Maddock, Laura D Howe, Eleanor M Winpenny

## Abstract

**Background:** Diet is an important risk factor for cardiovascular disease and shows well-established socioeconomic patterning among adults. However, less clear is how socioeconomic inequalities in diet develop across the life course. This study assessed the associations of early adulthood socioeconomic trajectories (SETs) with adult diet quality, adjusting for childhood socioeconomic position (SEP) and testing for mediation by adulthood SEP.

**Methods:** Participants from the 1970 British Cohort Study with socioeconomic data in early adulthood were included (n=12434). Diet quality at age 46 years, evaluated using the Mediterranean diet pyramid, was regressed on six previously identified classes of early adulthood SETs between ages 16 and 24 years, including a Continued Education class, four occupation-defined classes, and an Economically Inactive class. Causal mediation analyses tested the mediation of the association via household income and neighbourhood deprivation at age 46 years separately. Models were adjusted for sex, childhood SEP, adolescent diet quality and adolescent health.

**Results:** The Continued Education class showed the best diet quality at age 46 years, while little difference in diet quality was found among the remaining SET classes. The association between the Continued Education class and adult diet quality was independent of parental SEP in childhood, and was largely not mediated by household income or neighbourhood deprivation (0.7% and 3.7% of the total effect mediated, respectively) in mid-adulthood.

**Conclusions:** Early adulthood SETs independently contribute to adult diet quality, with continuing education associated with better adherence to Mediterranean diet. Early adulthood therefore represents a critical period for intervention to alleviate dietary inequalities in later life.

**Key Messages:** Life course epidemiology research aims to identify potential windows of change in life to prevent the establishment of socioeconomic inequalities in diet and diet-related cardiovascular health.

Early adulthood socioeconomic trajectories contributed to adult diet quality independent of childhood and adulthood socioeconomic position, with continuing education between ages 16 and 24 years associated with better diet quality at age 46 years.

Early adulthood represents a critical period for intervention to alleviate socioeconomic inequalities in diet persisting into adulthood.

## Introduction

Poor diet quality is the leading modifiable risk factor for cardiovascular disease and associated health inequalities in high-income countries.^1,2^ In the UK, particularly, there is a recent trend of widened inequalities in adult cardiovascular health,^3^ to which the well-established socioeconomic gradient in diet could contribute. Much research has shown that adults in low socioeconomic position (SEP) have low intakes of fruits, vegetables, whole grains and other healthy food items, but overconsume energy-dense and nutrient-poor foods,^4-7^ indicating a suboptimal diet quality.

However, it is less clear during which stage of life the socioeconomic gradient in diet is established. Understanding dietary inequalities from a life-course perspective is needed to identify target timings and populations for intervention to promote a healthy diet for all. Diet-related socioeconomic inequalities may start early in life. Low parental education was found consistently associated with worse diet quality among children,^8-10^ and this association was also observed but less consistent among adolescents.^11,12^ Moreover, dietary inequalities established in early life to some extent persist into adulthood. Results from life course epidemiology research show that parental SEP during childhood contributes to adult diet quality, independent of adulthood SEP.^13-15^

This study focused on another important but largely overlooked stage of life – early adulthood – in the development of socioeconomic inequality in diet. Early adulthood (ages 16-24 years) is a demographically dense period when young people go through multiple education- and employment-related life transitions.^16^ These education/employment transitions lead to the instability of SEP, and allow intergenerational social mobility between young people and their parents.^17,18^ The SEP established in early adulthood may further determine how much people earn and what type of neighbourhoods they live in later adulthood, which have previously shown associations with adult diet quality.^4,19,20^ Therefore, early adulthood may represent a critical stage of life for developing long-term diet and dietary inequalities.

In this study, we aim to understand the contribution of early adulthood SEP to adult diet quality from a life course perspective. A trajectory-based measure of early adulthood SEP was used to encompass changes in educational and economic activities from age 16 to 24 years, and to test associations with adult diet quality. Specifically, two research questions were included in this study:

1. Do socioeconomic trajectories (SETs) across early adulthood contribute to diet quality in mid-adulthood (age 46 years), independent of childhood SEP (age 10 years)?
2. To what extent does adulthood SEP (age 46 years) mediate the association between early adulthood SETs and adult diet quality?

## Methods

### Data

The data were from the 1970 British Cohort Study (BCS70).^21^ BCS70 recruited around 17,000 participants born in England, Scotland and Wales in one week of 1970, and has followed their lives across ten sweeps. Since age 16 years, participants had reported their main educational and economic activities for each year. This study included participants with data on educational and economic activities during early adulthood (ages 16-24 years, n=12434).

### Exposure

Early adulthood socioeconomic trajectories (SETs) were identified previously using data on educational and economic activities between ages 16 and 24 years.^22^ Based on year-specific activity data, a longitudinal latent class analysis generated six classes of SETs in early adulthood: (1) Continued Education, (2) Managerial Employment, (3) Skilled Non-manual Employment, (4) Skilled Manual Employment, (5) Partly Skilled Employment, and (6) Economically Inactive. Supplementary Figure S1 illustrates the activity participation with age, by early adulthood SET class. For example, the Continued Education class is dominated by participants who continued education after age 16 years and worked in a professional, managerial or skilled non-manual occupation at age 24 years. Compared to SEP measured at a single time point, the trajectory-based SEP measure can identify changes in two commonly used SEP indicators (occupational social class and length of participation in education) across early adulthood.

### Outcome

Data on dietary intake were collected using a web-based dietary intake questionnaire, the Oxford WebQ, at age 46 years. The Oxford WebQ was developed for monitoring diet and dietary changes in large-scale prospective studies, including questions on the consumption of 206 foods and 32 beverages in 24 hours. ^23,24^ Participants recorded the foods and drinks that they had consumed on the previous day. For the evaluation of diet quality independent of the total diet quantity, we uniformly adjusted energy intake to 2500 kcal/day (10.46 MJ/day), using the residual method (see supplementary methods for detail).^25^

The pyramid-based Mediterranean diet score (PyrMDS) was used as the measure of overall diet quality. PyrMDS accounts for both traditional Mediterranean diet and the contemporary food environment,^26^ and adherence to it has shown associations with a reduced risk of dementia, cardiovascular diseases and all-cause mortality.^27-29^ The algorithm for PyrMDS includes fifteen food components, with each component assigned a continuous score from 0 to 1 based on recommended daily consumption.^30^ One exception is ‘olive oil’, which we assigned a discrete score of 1 or 0 to indicate consumers (3.5% of the participants) or non-consumer, respectively. PyrMDS was calculated by summing the scores of fifteen food components (range: 0-15), with a higher score representing greater adherence to the Mediterranean diet pyramid. Table S1 provides the food items and components included and the method for calculation of PyrMDS.

### Covariates

Four groups of covariates that could confound the association between early adulthood SETs and adult diet quality were included in the analysis: (1) sex, (2) childhood SEP, (3) adolescent diet quality, and (4) adolescent health. Testing for the interaction of sex and early adulthood SETs did not find effect moderation by sex. Childhood SEP included measures of parental education, parental social class, family income, family structure and neighbourhood location at age 10 years, and self-reported social rating of the neighbourhood at age 5 years. Adolescent diet quality was recorded using a diet diary at age 16 years and evaluated using energy-adjusted PyrMDS, excluding the components of wine (not recommended for adolescents) and fat (olive oil unavailable from the dietary data). Adolescent health included measures of health-related behaviours and health status at age 16 years. Further details of all covariates are provided in Table S2.

### Mediators

Household and neighbourhood SEP at age 46 years were tested as potential mediators of the association between early adulthood SETs and adult diet quality. For household SEP, equivalised household income was calculated as weekly household income divided by (1 + 0.5 x number of additional adults + 0.3 x number of children aged 0-15 years).^31^ For neighbourhood SEP, neighbourhood deprivation was measured by the Index of Multiple Deprivation Score.^32^ The deprivation index is a composite measure of relative deprivation based on specific life domains for each lower super output area in England (2015) and Wales (2014) and for each data zone in Scotland (2016). Relative deprivation of the neighbourhood was assessed by decile values, with 1 representing the most deprived area and 10 the least deprived area.

### Statistical analysis

Descriptive and modelling analyses were conducted in R. We compared diet quality at age 46 years among six classes of early adulthood SETs, based on the mean and standard deviation (SD) of PyrMDS and the consumption of Mediterranean diet components. Independent t-tests (for continuous diet measures) and chi-square tests (for binary diet measures) were used to examine dietary differences between the Continued Education class and the remaining SET classes. As all participants who had data on early adulthood SETs were included (n=12423), we imputed missing covariate and outcome data using multiple imputation by chained equations with auxiliary variables (see Table S2 for detail).^33^

Ordinary least squares (OLS) regression was performed to test the overall association between six classes of early adulthood SETs and adult diet quality. We used the *marginal_means* command in the *marginaleffects* R package to calculate the predicted mean of PyrMDS for each SET class, after adjusting for covariates. Given the negligible difference in the predicted PyrMDS among the four occupation-defined SET classes and the Economically Inactive class (see OLS regression results below), we dichotomised early adulthood SET classes into the Continued Education classes and the remaining SET classes (as the reference category) for the mediation analysis.

The causal mediation analysis, using *mediation* R package, assessed the mediating pathway from early adulthood SETs to adult diet quality via adult SEP. Household income and neighbourhood deprivation were tested as the mediator separately, since both of them were measured at age 46 years and could be regarded as causally unrelated mediators.^34,35^ The causal mediation analysis draws upon a counterfactual framework to decompose the total effect into average direct effect and average causal mediation effect (see supplementary methods for the effect estimation methods).^35^ For example, average causal mediating effect evaluated how the predicted PyrMDS would change when the mediator took the predicted value at the Continued Education class versus at another SET class, while the assignation to a SET class remained unchanged. The proportion mediated was computed as the ratio between the average causal mediation effect and the total effect. The interaction terms of the Continued Education class and two mediators were also included to test for potential interaction effects. To examine the robustness of the estimated mediation effects, we conducted two supplementary analyses respectively on the bias from mediator-outcome confounders and the exclusion of the Economically Inactive class (see supplementary methods for details of the sensitivity analysis).

## Results

### Participants’ socioeconomic characteristics

Table 1 shows descriptive statistics of the BCS70 participants (n=12423) by early adulthood SET class. The Continued Education class included a higher proportion of participants whose parents were in the professional and managerial social class and achieved a university/higher level of education than the remaining SET classes. Participants from the Continued Education class also had the highest equivalised household income and resided in a neighbourhood with the lowest deprivation levels on average at age 46 years, suggesting tracking of SEP from childhood to adulthood.

**Table 1.**
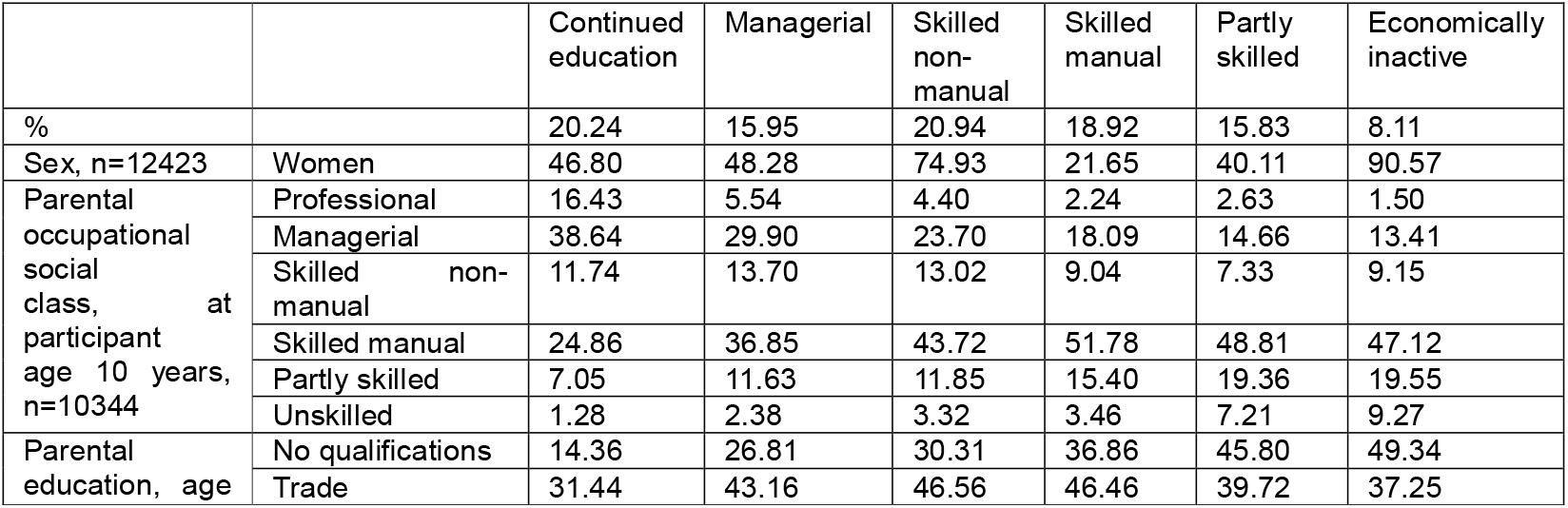
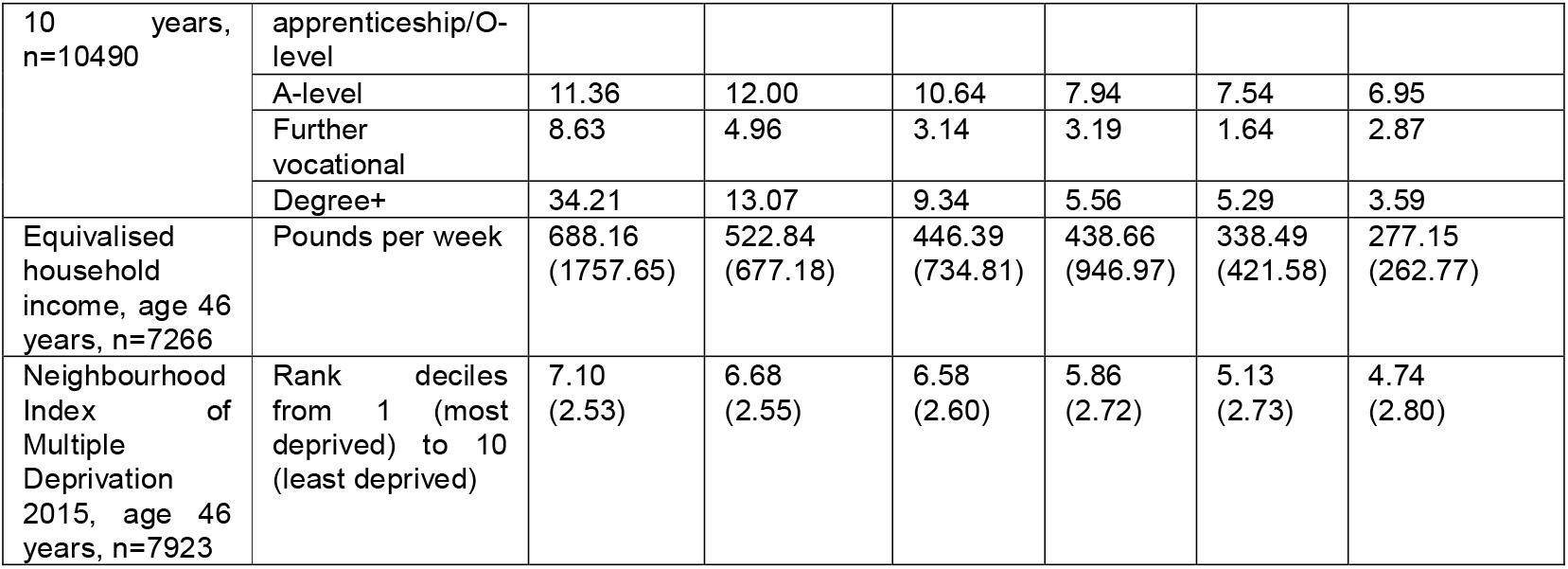
Descriptive statistics of the BCS70 participants by early adulthood SET (n=12423)

### Diet quality at age 46 years by early adulthood socioeconomic trajectory

Participants from the Continued Education class showed better diet quality at age 46 years, as suggested by higher mean values of PyrMDS and mean intakes of most food components more aligned with the Mediterranean diet (except for legumes, white meat and eggs), compared to the other five SET classes (Table 2).

**Table 2.**
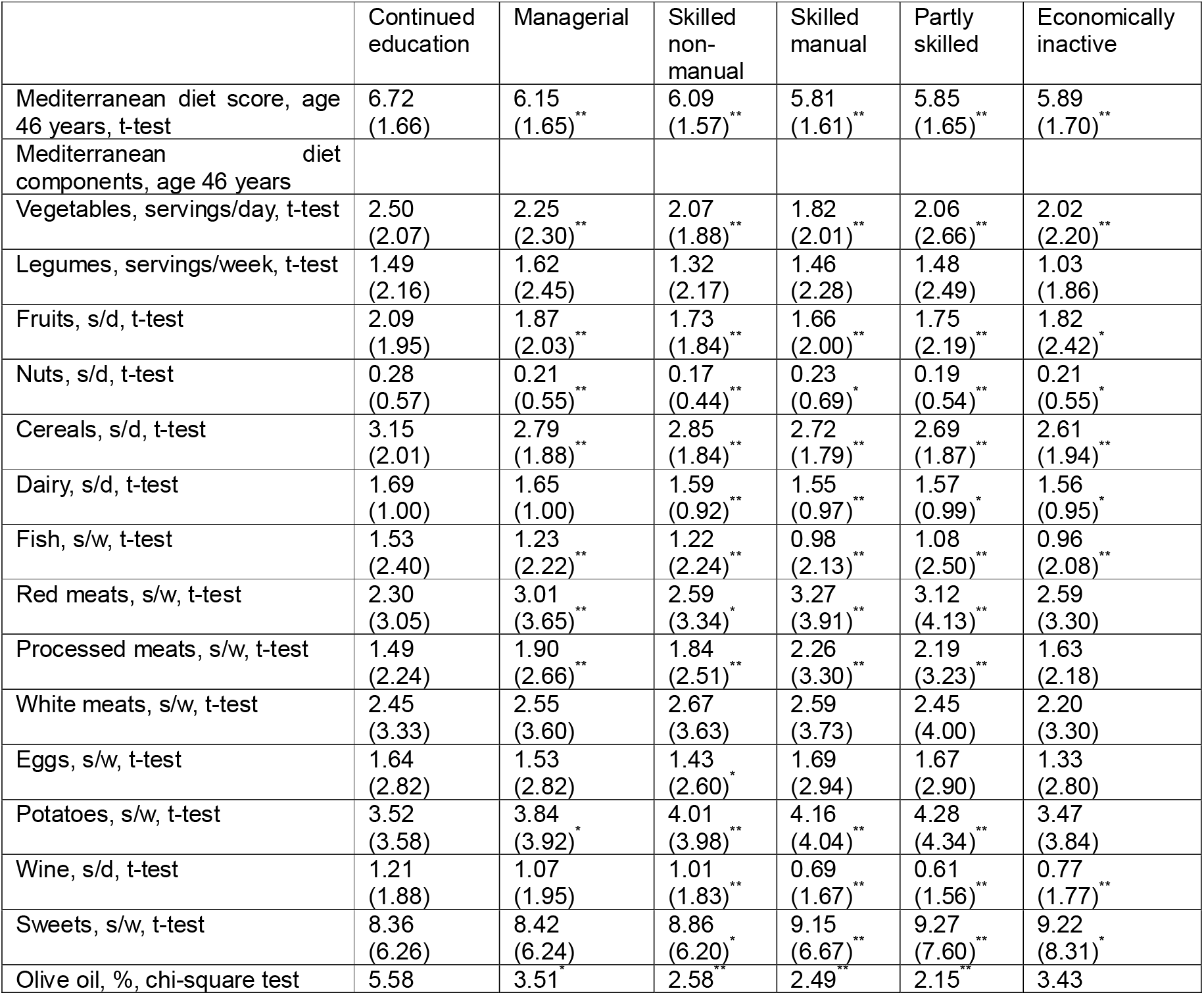
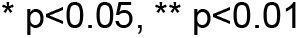
Mediterranean diet scores and components at age 46 years by early adulthood SET.

|  | Continued education | Managerial | Skilled non-manual | Skilled manual | Partly skilled | Economically inactive |
| --- | --- | --- | --- | --- | --- | --- |
| Mediterranean diet score, age 46 years, t-test | 6.72<br>(1.66) | 6.15<br>(1.65)** | 6.09<br>(1.57)** | 5.81<br>(1.61)** | 5.85<br>(1.65)** | 5.89<br>(1.70)** |
| Mediterranean diet components, age 46 years |  |  |  |  |  |  |
| Vegetables, servings/day, t-test | 2.50<br>(2.07) | 2.25<br>(2.30)** | 2.07<br>(1.88)** | 1.82<br>(2.01)** | 2.06<br>(2.66)** | 2.02<br>(2.20)** |
| Legumes, servings/week, t-test | 1.49<br>(2.16) | 1.62<br>(2.45) | 1.32<br>(2.17) | 1.46<br>(2.28) | 1.48<br>(2.49) | 1.03<br>(1.86) |
| Fruits, s/d, t-test | 2.09<br>(1.95) | 1.87<br>(2.03)** | 1.73<br>(1.84)** | 1.66<br>(2.00)** | 1.75<br>(2.19)** | 1.82<br>(2.42)* |
| Nuts, s/d, t-test | 0.28<br>(0.57) | 0.21<br>(0.55)** | 0.17<br>(0.44)** | 0.23<br>(0.69)* | 0.19<br>(0.54)** | 0.21<br>(0.55)* |
| Cereals, s/d, t-test | 3.15<br>(2.01) | 2.79<br>(1.88)** | 2.85<br>(1.84)** | 2.72<br>(1.79)** | 2.69<br>(1.87)** | 2.61<br>(1.94)** |
| Dairy, s/d, t-test | 1.69<br>(1.00) | 1.65<br>(1.00) | 1.59<br>(0.92)** | 1.55<br>(0.97)** | 1.57<br>(0.99)* | 1.56<br>(0.95)* |
| Fish, s/w, t-test | 1.53<br>(2.40) | 1.23<br>(2.22)** | 1.22<br>(2.24)** | 0.98<br>(2.13)** | 1.08<br>(2.50)** | 0.96<br>(2.08)** |
| Red meats, s/w, t-test | 2.30<br>(3.05) | 3.01<br>(3.65)** | 2.59<br>(3.34)* | 3.27<br>(3.91)** | 3.12<br>(4.13)** | 2.59<br>(3.30) |
| Processed meats, s/w, t-test | 1.49<br>(2.24) | 1.90<br>(2.66)** | 1.84<br>(2.51)** | 2.26<br>(3.30)** | 2.19<br>(3.23)** | 1.63<br>(2.18) |
| White meats, s/w, t-test | 2.45<br>(3.33) | 2.55<br>(3.60) | 2.67<br>(3.63) | 2.59<br>(3.73) | 2.45<br>(4.00) | 2.20<br>(3.30) |
| Eggs, s/w, t-test | 1.64<br>(2.82) | 1.53<br>(2.82) | 1.43<br>(2.60)* | 1.69<br>(2.94) | 1.67<br>(2.90) | 1.33<br>(2.80) |
| Potatoes, s/w, t-test | 3.52<br>(3.58) | 3.84<br>(3.92)* | 4.01<br>(3.98)** | 4.16<br>(4.04)** | 4.28<br>(4.34)** | 3.47<br>(3.84) |
| Wine, s/d, t-test | 1.21<br>(1.88) | 1.07<br>(1.95) | 1.01<br>(1.83)** | 0.69<br>(1.67)** | 0.61<br>(1.56)** | 0.77<br>(1.77)** |
| Sweets, s/w, t-test | 8.36<br>(6.26) | 8.42<br>(6.24) | 8.86<br>(6.20)* | 9.15<br>(6.67)** | 9.27<br>(7.60)** | 9.22<br>(8.31)* |
| Olive oil, %, chi-square test | 5.58 | 3.51 | 2.58 | 2.49 | 2.15 | 3.43 |

\* $p < 0.05$ , \*\* $p < 0.01$

### The association between early adulthood socioeconomic trajectories and adult diet quality

Figure 1 illustrates estimated marginal means of PyrMDS at age 46 years by early adulthood SET, after adjusting for covariates (numerical results for the OLS regression model presented in Table S3). The modelled mean value of PyrMDS was significantly higher for the Continued Education class than the remaining SET classes. Across the remaining SET classes, there were slight downward gradient in the mean values of PyrMDS based on the ranking of occupational social class.

**Figure 1.**
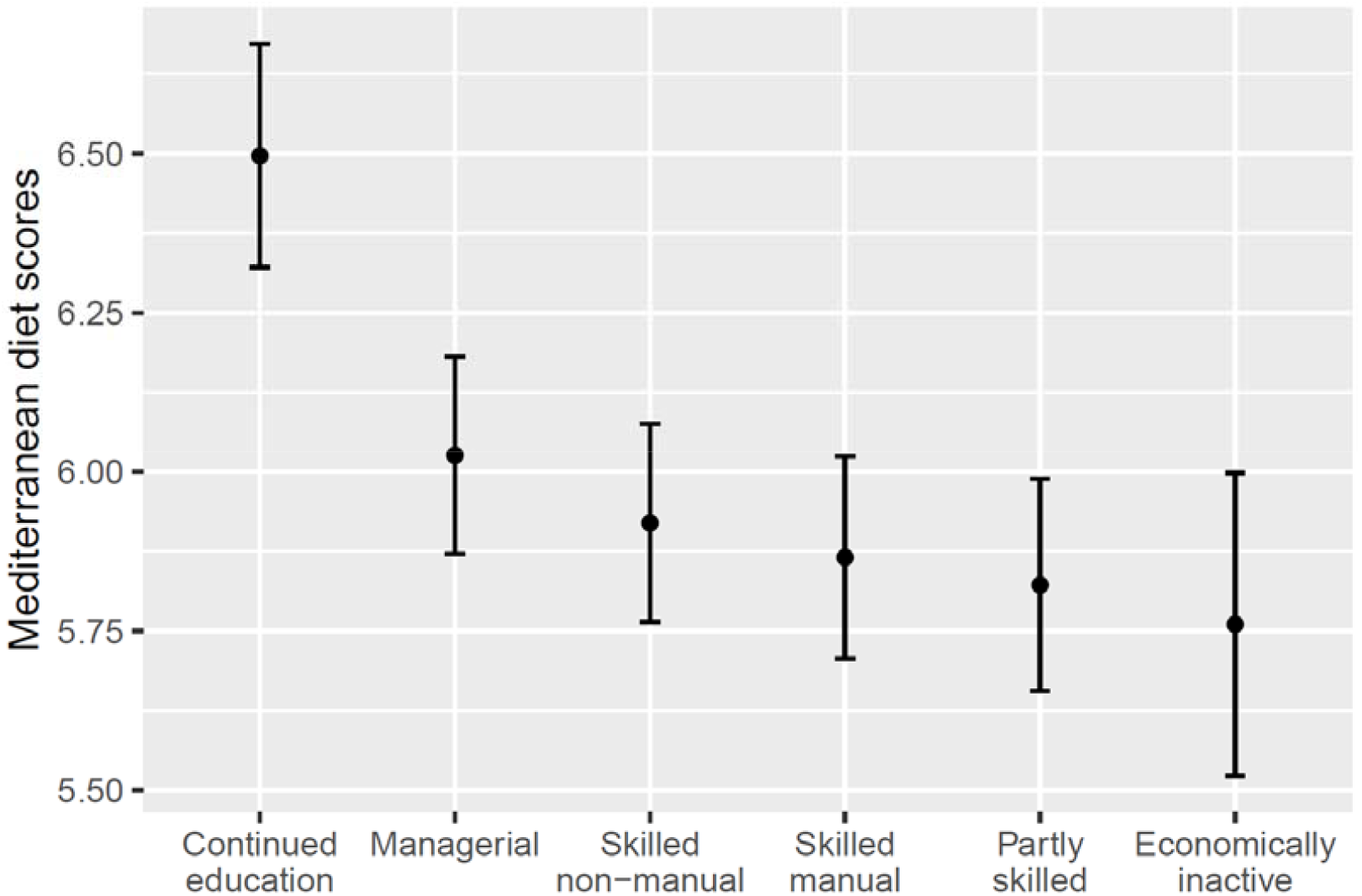
Modelled mean values (with 95% CIs) of Mediterranean diet scores at age 46 years by early adulthood SET Results are adjusted for the covariates of sex, childhood SEP (age 10 years), baseline Mediterranean diet (age 16 years), and baseline health-related behaviours and health status (age 16 years).

### Mediation of the association by household and neighbourhood socioeconomic position

Figure 2 demonstrates the mediation of household and neighbourhood SEP in the association between the Continued Education class and adult diet quality (with the remaining SET classes as the reference). Results from causal mediation models suggested household income did not mediate the association (0.7% of the proportion mediated). Weak mediation was found for neighbourhood deprivation (3.7% of the proportion mediated). Participants of the Continued Education class were more likely to reside in less deprived neighbourhoods at age 46 years, while lower levels of neighbourhood deprivation were weakly associated with better diet quality. For both mediators, little evidence of exposure-mediator interaction was observed.

**Figure 2.**
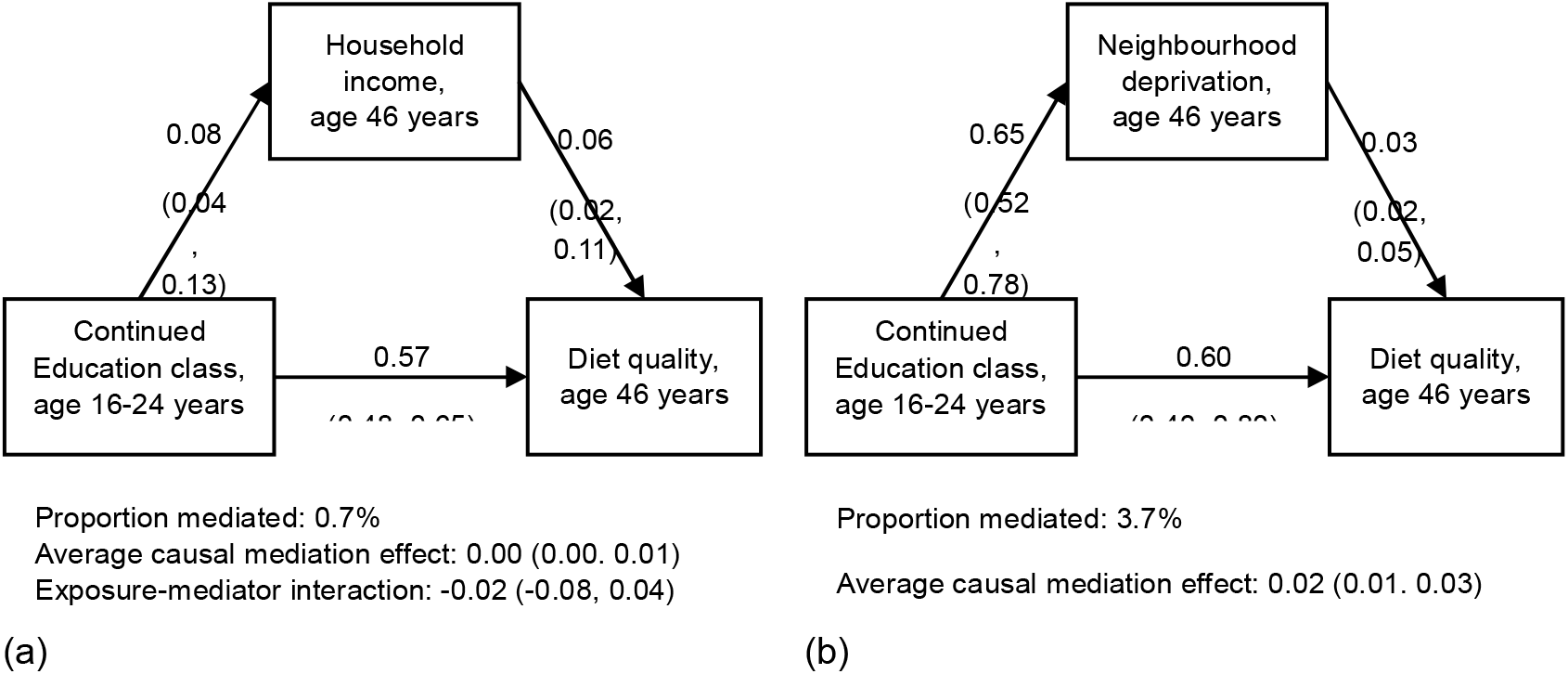
The causal mediation analysis of the association between early adulthood socioeconomic trajectories and adult diet quality via (a) household income and (b) neighbourhood deprivation at age 46 years Results are shown in β (95% CI), and are adjusted for covariates. Results from the direct effects are presented on each arrow of the diagrams.

The mediation of neighbourhood deprivation was sensitive to the existence of uncontrolled mediator-outcome confounders, since the 95% CI of the estimated mediation effect would span the null as we increased the correlation between the residuals of the mediator and outcome regressions (Figure S2). After excluding the Economically Inactive class from the causal mediation analysis, we found little change in the mediation effects of household and neighbourhood SEP, except an attenuated association between household income and diet quality (n=11416; Figure S3).

## Discussion

### Main findings from this study

This study examined the contribution of the transitional period of early adulthood (ages 16-24 years) to socioeconomic inequalities in adult diet quality. Using a trajectory-based measure of SEP across early adulthood, we found that the Continued Education class was associated with better adherence to the pyramid-based Mediterranean diet at age 46 years, compared to the remaining classes. This SET-diet association was largely not mediated by household income or neighbourhood deprivation in mid-adulthood, and was independent of parental and neighbourhood SEP during childhood.

### Comparison with previous research

Few studies have focused on the period of early adulthood when investigating socioeconomic inequalities in diet. Some studies even excluded adult participants aged below 25 years because of unstable socioeconomic circumstances during this period.^36-38^ A recent study on within-person dietary changes across early adulthood transitions shows that leaving early from full-time education was associated with persistent decreases in diet quality across early adulthood, while starting a first job at an older age of early adulthood was related to more beneficial dietary changes.^39^ This study develops the evidence by demonstrating a longer-term association between continuing education across early adulthood and better diet quality in mid-adulthood. There was little difference in diet quality across four occupation-defined SET classes and a larger variance of diet quality for the Economically Inactive class. Consistent with other studies,^40,41^ education represents a more reliable indicator in early adulthood than occupational social class to represent enduring socioeconomic circumstances in later life.

The association between early adulthood SETs and adult diet quality was established after adjustment for childhood SEP. This is consistent with previous cross-sectional studies, where parental education levels during childhood were associated with participants’ dietary intake, but participants’ own educational attainment played a more influential role, or at least attenuated the relationship between parental education and adult diet quality.^13-15,36,42^ A more direct test of social mobility effects also shows that participants whose education levels were lower than their parents reported lower intake of fruits and vegetables and worse overall diet quality than participants as well-educated as their parents,^41,43^ indicating an independent contribution of participants’ own education.

Little mediation by household income was observed of the association between the Continued Education class and adult diet quality. Previous studies suggest that education and income reflect different dimensions of the socioeconomic construct. Educational attainment, an indicator of the acquisition of knowledge and skills, showed more consistent associations with diet outcomes compared to the material resources acquired from income.^37,42,44^ A cross-sectional study of the UK adult population observed a partial mediation of income in the relationship between education and diet quality.^19^ Our longitudinal analysis made clear the temporal order between SEP indicators, and found negligible mediation of the association between continuing education and adult diet quality via household income at age 46 years.

In contrast, there was slight mediation of the SET-diet association by neighbourhood deprivation. Previous research findings are mixed for the relationship between living in socioeconomically deprived neighbourhoods and low intake of fruits and vegetables in high-income countries.^45^ In the US, unhealthy food environments were disproportionally distributed in low-income and racial-minority neighbourhoods, resulting in worse diet quality for residents living there.^4,20^ However, this deprivation amplification hypothesis was largely not substantiated in other high-income countries.^41,46,47^ In the UK, particularly, more deprived neighbourhoods could provide convenient access to supermarkets and grocery stores with healthy food choices, despite their greater exposure to fast-food outlets than less deprived neighbourhoods.^5,48,49^

### Strengths and limitations

A major strength of this study is the use of a life course approach, following participants’ socioeconomic circumstances from childhood to adulthood, to understand the development of socioeconomic inequalities in overall diet quality in mid-adulthood. We drew upon a longitudinal trajectory-based measure of SEP to delineate the dynamics in educational participation and occupational social class across early adulthood. Moreover, the causal mediation analysis found little mediation of the SET-diet relationship via adulthood SEP. It is therefore still unclear the pathways underlying the contribution of early adulthood SETs to adult diet quality. Due to unavailability of longitudinal diet data, we cannot test the pathway of tracking of behaviours, where the dietary patterns established in early adulthood may persist into adulthood and contribute to adult dietary inequalities.^16,39^

The BCS70 study recruited a large birth cohort born in 1970 in the UK. To alleviate attrition bias over study sweeps, we included participants who reported educational and economic activity data in early adulthood, imputed missing outcome data, and controlled for childhood covariates including those associated with attrition. The measure of early adulthood SETs provides an overview of unstable socioeconomic circumstances experienced in this period, but is not comparable to educational achievement or occupational status assessed at a single point of time in previous studies. The classes of early adulthood SETs for the 1970 cohort may not be representative of a younger generation, which needs updated longitudinal analysis based on more recent birth cohorts.

This study relied on a validated 24-hour food record to comprehensively assess dietary intake at age 46 years.^24^ The 24-hour food record, however, is unlikely to represent individual’s habitual diet, but has been shown to adequately reflect group-level intake across population groups.^50^ Participants reported their food intake with error, so we uniformly adjusted the energy consumption to evaluate diet quality independent of diet quantity. However, other systematic biases may remain. For example, it is unclear if certain socioeconomic groups are more likely to underreport foods considered to be less healthy and overreport those considered to be more healthy, thereby causing biased estimations of socioeconomic inequalities in diet.

## Conclusions

Life course epidemiology research aims to identify potential windows of change in life to prevent the establishment of socioeconomic inequalities in diet and diet-related cardiovascular health.^7,12,17^ This study highlights that early adulthood SETs contribute to adult diet quality independent of childhood and adulthood SEP, with continuing education between ages 16 and 24 years associated with better diet quality at age 46 years. Early adulthood therefore represents a critical period for intervention to alleviate dietary inequalities in later life (Figure 3). Given little mediation of the association between early adulthood SETs and adult diet quality via adulthood SEP, further research needs to examine other pathways underlying the association from a life-course perspective, including cumulative exposures to food environments and tracking of dietary patterns with age.

**Figure 3.**
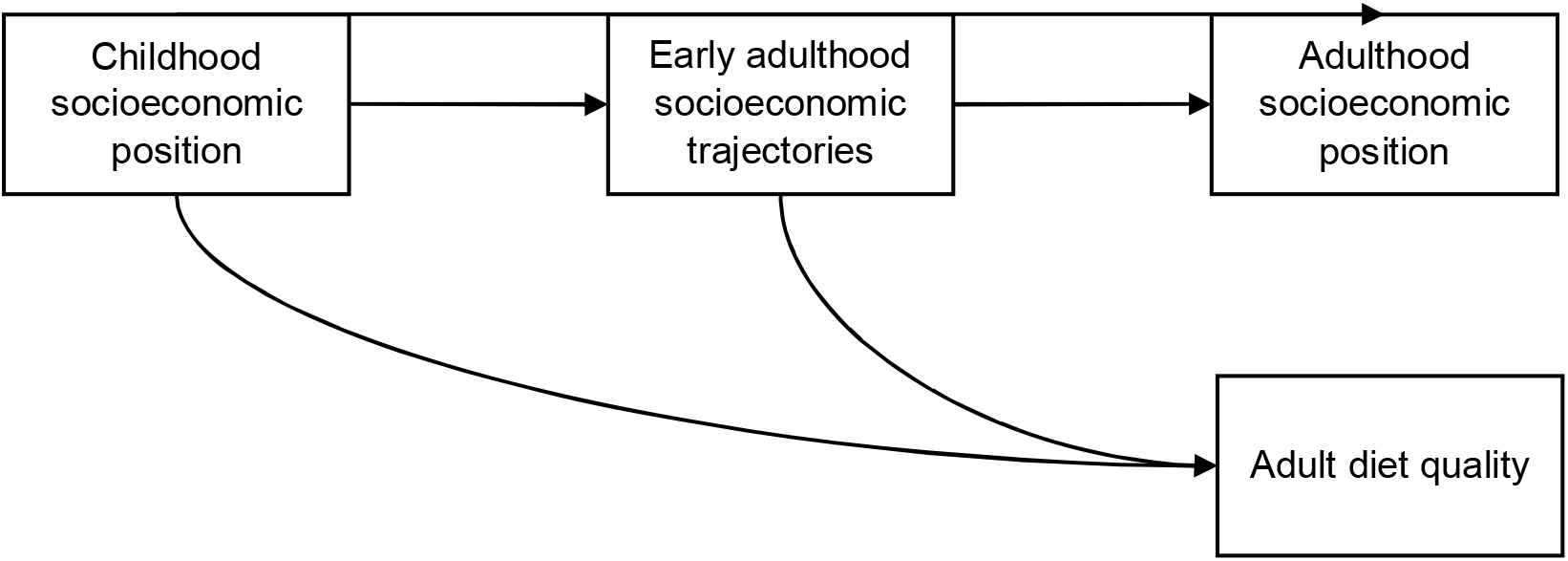
A life course perspective for understanding socioeconomic inequality in diet quality

## Data Availability

Data are available in a public, open access repository. The data underlying this article are freely available to bona fide researchers via the UK Data Service (http://ukdataservice.ac.uk).

## Ethics approval

Ethical approval for the age 46 follow--up of the 1970 British Cohort Study was obtained from the South East Coast–Brighton and Sussex National Research Ethics Service (NRES) Committee (Ref. No. 15/LO/1446). Previous waves of the study were approved by other NRES committees (year 2000 onwards) or local committees. This research was conducted in accordance with the ethical standards laid down in the 1964 Declaration of Helsinki and its later amendments.

## Supplementary Data

Supplementary data are available at IJE online.

## Author contributions

YT and EMW designed the research, YT conducted the analyses and drafted the manuscript, EMW, JM and LDH revised the manuscript. All authors commented on the analysis plan, contributed to the interpretation of results and critically reviewed the final manuscript. The author(s) read and approved the final manuscript.

## Acknowledgements

The authors would like to thank all participants of the 1970 British Birth Cohort, as well as everyone who contributed to the data collection.

