## Supplementary documents for "Early adulthood socioeconomic trajectories contribute to inequalities in adult diet quality, independent of childhood and adulthood socioeconomic position"

**Supplementary information**

**Supplementary methods**

Energy adjustment for food intake: further details of the residual method

For the evaluation of diet quality independent of the total diet quantity, we uniformly adjusted energy intake to 2500 kcal/day (10.46 MJ/day), using the residual method. In this method, the energy-adjusted food intake is the residual from the regression model in which total energy intake in the survey day is the independent variable and absolute food intake is the dependent variable. The residual is therefore an estimate of food intake uncorrelated with total energy intake and directly related to overall variation in food choice and composition. The estimated terms in the regression model (formula 1) are used to calculate the predicted food intake with total energy intake set at 2500 kcal/day.

$EFI= \alpha+\beta E+ \sigma$	(1),

in which $EFI$ represents energy-adjusted food intake, $E$represents total energy intake, $\alpha$, $\beta$ and $\sigma$ are respectively the constant term, coefficient and residual error of the regression model.

Causal mediation analysis: further details of the effect estimation

In the causal mediation analysis, average direct effect (ADE; formula 1) evaluated the difference in the predicted value of PyrMDS when a participant switched from the Continued Education class to another SET class, while the mediator, household income or neighbourhood deprivation, was held constant at the predicted value that would take for a given SET class. Average causal mediating effect (ACME; formula 2) evaluated how the predicted value of PyrMDS would change when the mediator took the predicted value at the Continued Education class versus at another SET class, while a participant’s assignation to a SET class remained unchanged.

$$ADE= Y_{i}\left( 1, M_{i}\left( x \right) \right)- Y_{i}\left( 0, M_{i}\left( x \right) \right) \left( 1 \right)$$

$$ACME= Y_{i}\left( x, M_{i}\left( 1 \right) \right)- Y_{i}\left( x, M_{i}\left( 0 \right) \right) \left( 2 \right),$$

where $M_{i}\left( x \right)$ represents the predicted value of a mediator $M$ for unit $i$ under the exposure status $x$, and $Y_{i}\left( x, M_{i}\left( x \right) \right)$ represents the predicted value of the outcome that would result if the exposure and mediator take the exposure status $x$ (1 as the treatment category and 0 as the reference category).

Causal mediation analysis: further details of the sensitivity analysis

To examine the robustness of the estimated mediation effects, we conducted two supplementary analyses. First, we tested possible biases from mediator-outcome confounders. Based on the assumption of sequential ignorability (Imai et al., 2010), we varied the values of correlation coefficients between the residuals of the mediator and outcome regressions to observe changes in the estimated ACMEs. Second, we excluded participants of the Economically Inactive class from the mediation analysis, considering that this class was different from the remaining occupation-driven SET classes, and showed a large variance in the predicted value of PyrMDS in the OLS regression.

**Supplementary figures**

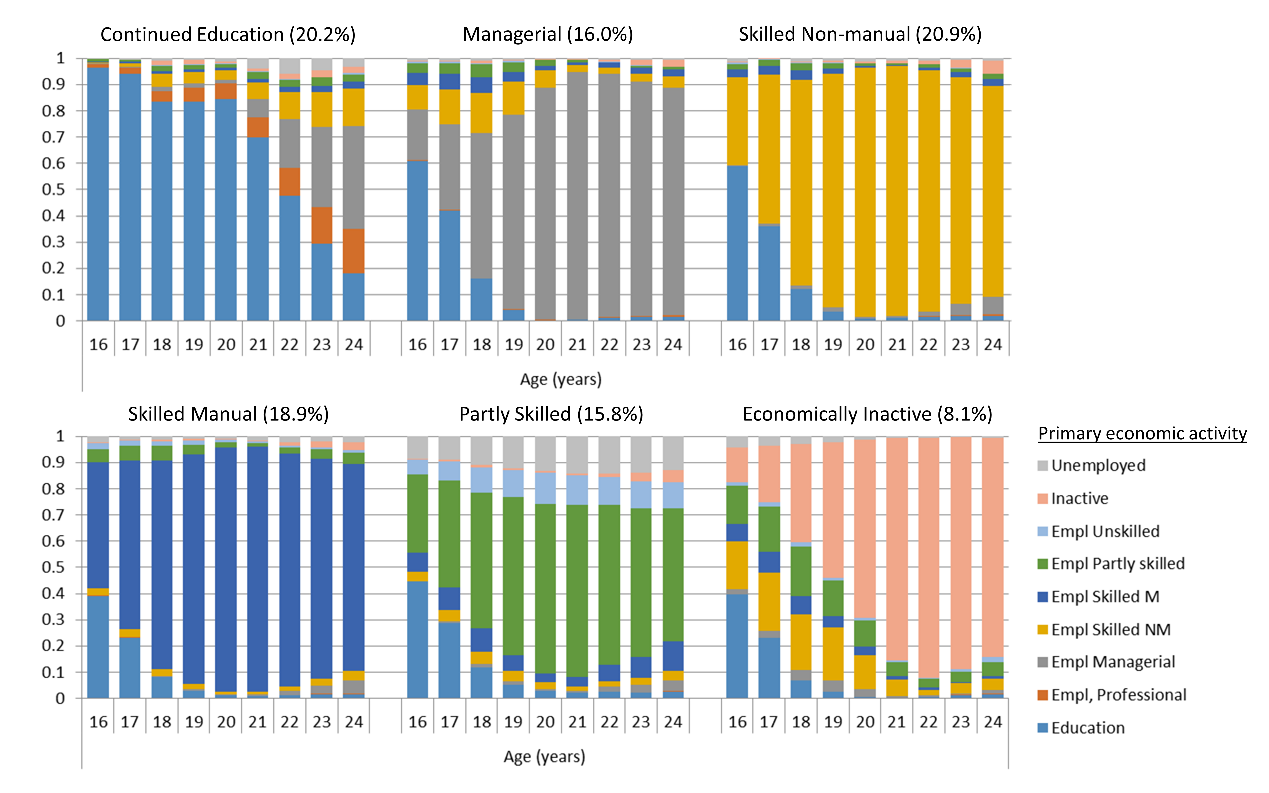

The six-class solution has an entropy of 0.97, indicating a high classification accuracy.

Figure S1. The six socioeconomic trajectory classes, showing response probabilities for participation in different educational and economic activities at each year of age (Winpenny et al., 2021)

| 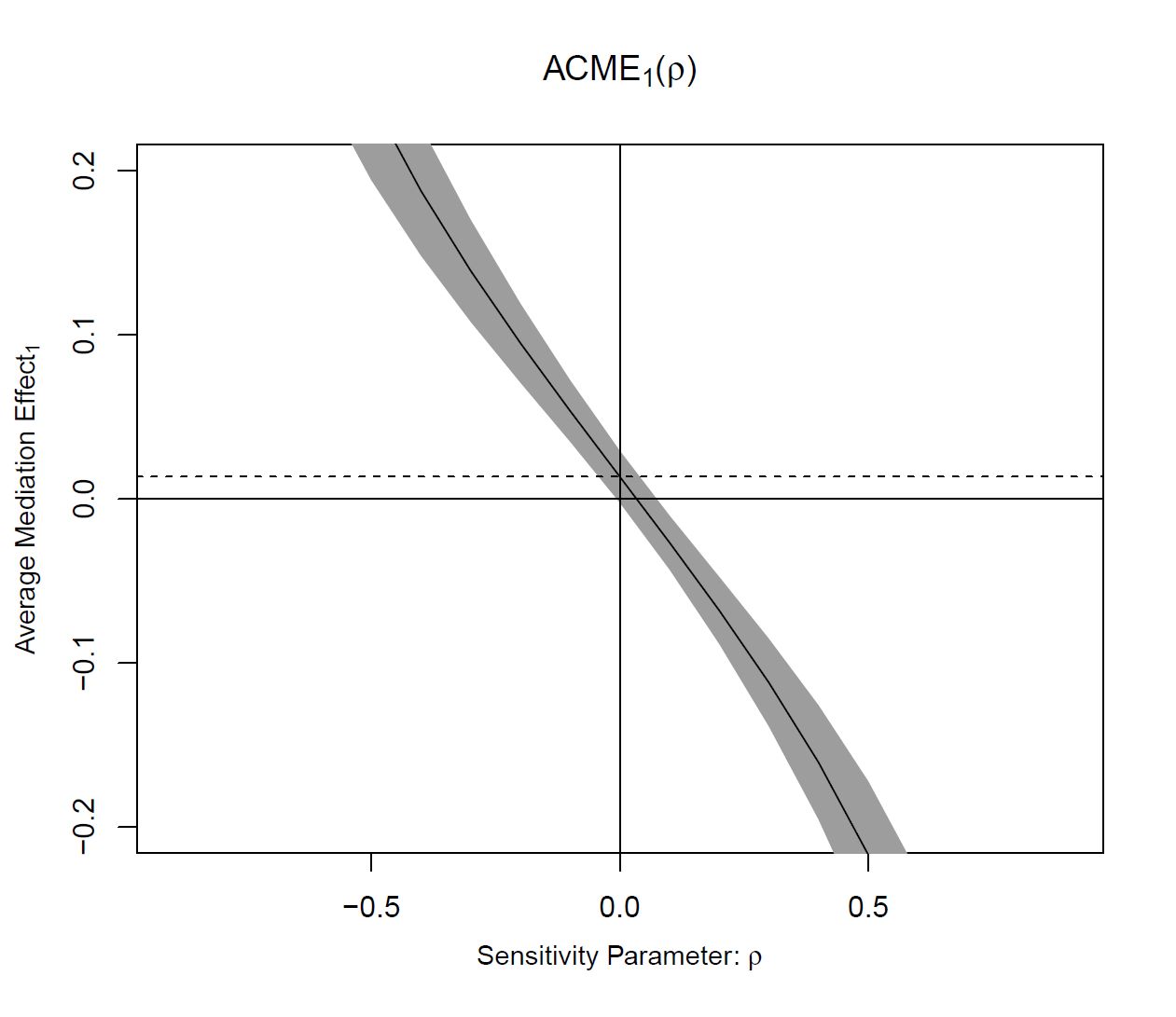 | 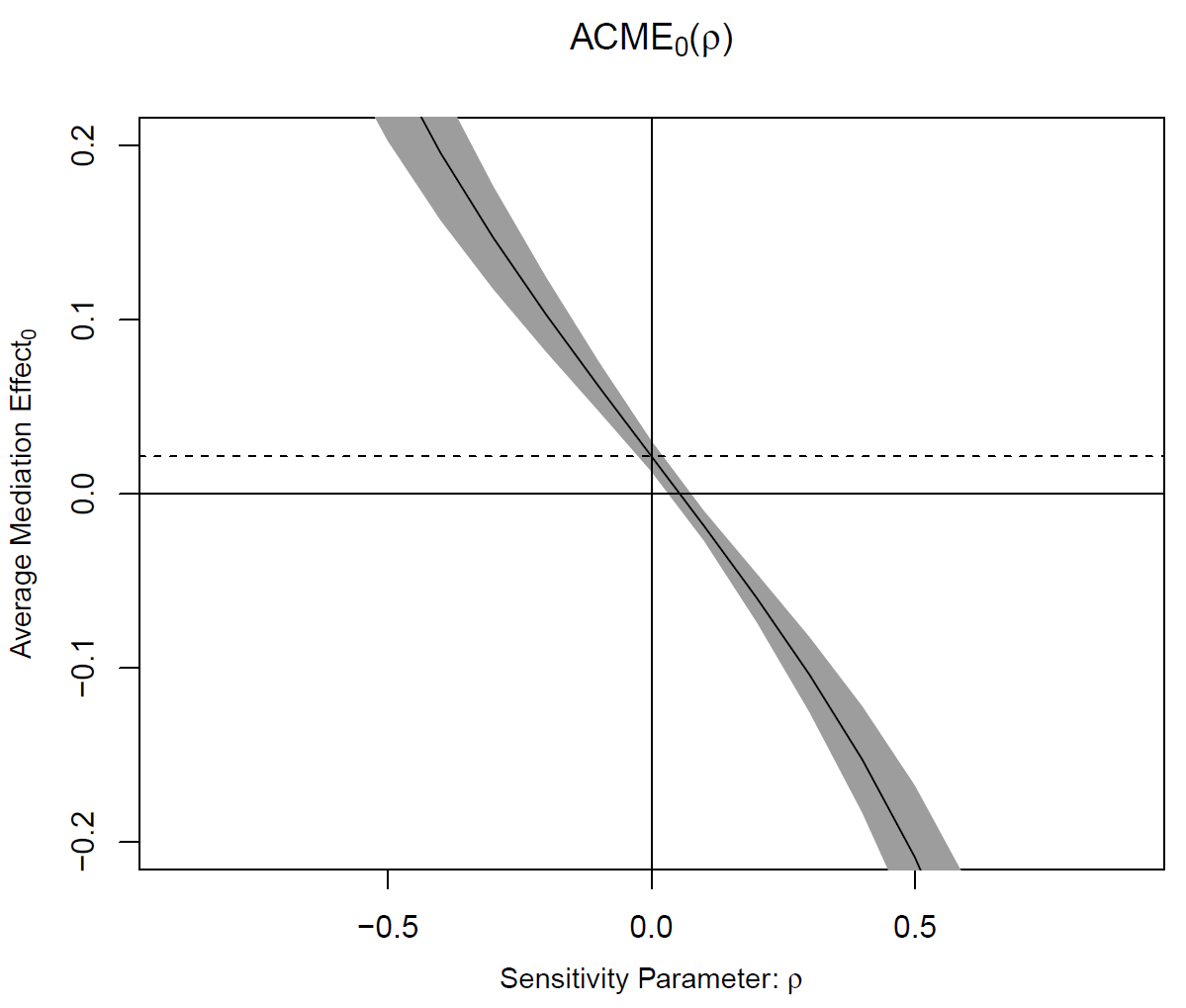 |
| --- | --- |
| (a) the Continued Education class | (b) the remaining SET classes |

Figure S2. A sensitivity test for the mediation of neighbourhood deprivation in the association between early adulthood socioeconomic trajectories and adult diet quality, as a function of the correlation between the residuals of the mediator and outcome regression models

| Continued Education class, ages 16-24  Household income,  age 46  Diet quality,  age 46  Proportion mediated: 0.4%  Average causal mediation effect: 0.00 (0.00. 0.01)  Exposure-mediator interaction: 0.06 (-0.01, 0.12)  0.05  (0.01, 0.10)  0.01  (-0.03, 0.05)  0.53  (0.44, 0.61) | Continued Education class, ages 16-24  Neighbourhood deprivation,  age 46  Diet quality,  age 46  Proportion mediated: 3.5%  Average causal mediation effect: 0.02 (0.01. 0.03)  Exposure-mediator interaction: 0.01 (-0.02, 0.03)  0.54  (0.41, 0.66)  0.03  (0.02, 0.05)  0.48  (0.29, 0.68) |
| --- | --- |
| (a) | (b) |

Results are shown in β (95% CI), and are adjusted for covariates.

Figure S3. The causal mediation analysis of the association between early adulthood socioeconomic trajectories and adult diet quality via (a) household income and (b) neighbourhood deprivation at age 46 years, after excluding the participants assigned to the Economically Inactive class (n=11416)

**Supplementary tables**

Table S1. The pyramid-based Mediterranean diet score (PyrMDS), food components and food items at age 16 and 46 years for the BCS70 participants

| **Food components** | **Food items at 16 years** | **Food items at 46 years** | **PyrMDS (0-15)** | |
| --- | --- | --- | --- | --- |
|  |  |  | Serving required for the score of 0 | Serving required for the score of 1 |
| Vegetables | fresh and frozen vegetables, canned and processed vegetables, tomatoes, green vegetables, carrots, salad vegetables | raw salad, green leafy/cabbages, root vegetables, tomatoes, allium vegetables, other vegetables (including mushrooms), fruiting and mixed vegetables, vegetable side dishes, vegetable dips | 0/d | ≥6/d |
| Legumes | peas, baked beans | meat substitutes – soy, peas/sweetcorn, legumes & pulses | 0/wk | ≥2/wk |
| Fruits | fresh fruit, not citrus/apple/pear, canned sweetened fruit, citrus fruits, apple | citrus, berries, apples & pears, other fruit, dried fruit, stewed fruit | 0/d | 3-6/d |
| Nuts | nuts | salted nuts & seeds, unsalted nuts & seeds | 0/d | 1-2/d |
| Cereals | white bread, wholemeal bread, brown wheatgerm granary bread, other breads, high fibre breakfast cereals, other breakfast cereals, unsweetened, sweetened breakfast cereals, pasta rice cereals | white bread, wholemeal bread, mixed (50/50), brown & seeded, other bread, bran cereal, biscuit cereal, oat cereal (non sugar), oat cereal (sugar), muesli, other cereal (sugar), white pasta & rice, wholemeal pasta, brown rice & other wholegrains, grain dishes - added fat | 0/d | 3-6/d |
| Dairy | whole milk, semi-skimmed milk, skimmed milk, other milk and cream, yoghurt, cottage cheese, cheese | whole milk, semiskimmed milk, skimmed milk, rice/oat milk, soy milk, full fat yogurt, low fat yogurt, high fat cheese, medium and low fat cheese | 0/d | 1.5-2.5/d |
| Fish | fish, fish dishes | white fish & tinned tuna, shellfish, oily fish, breaded/battered fish | 0/wk | ≥2/wk |
| Red meats | beef, lamb and mutton, pork, offal and dishes, meat dishes | pork, beef, lamb, other meat & offal | ≥4/wk | ˂2/wk |
| Processed meats | meat pies, burgers & kebabs, sausages, bacon & ham, canned meat | processed meat, breaded/battered chicken | ≥2/wk | ≤1/wk |
| White meats | coated chicken products | poultry | 0/wk | 1.5-2.5/wk |
| Eggs | eggs, egg and cheese dishes | egg & egg dishes | 0/wk | 2-4/wk |
| Potatoes | potatoes not fried, fried, roast potato not chips | potatoes/sweet potatoes (baked/boiled), mashed potatoes, fried/roast potatoes | ≥6/wk | ≤3/wk |
| Wine | – | white wine, red wine, fortified wine | ≥4/d for men,  ≥2/d for women | 1.5-2.5/d for men,  0.5-1.5/d for women |
| Sweets | sweet biscuits, cakes buns pastries, sugar confectionery, chocolate confectionery, ice cream lollies, puddings and fruit pies, milk puddings | added sugars & preserves, chocolate confectionery, other sweets, biscuits, milk-dairy desserts, other desserts & cakes & pastries | ≥4/wk | ≤2/wk |
| Fat | – | olive oil | Non-consumers | Consumers |

A moderate intake of fruits, nuts, cereals, dairy, white meat, and eggs was recommended. Points were allocated continuously between 0 (no consumption) and 1 (achieving an intake within the recommended level). Overconsumption (consuming an amount double the mid-point of the recommended intake), was penalised and received a maximum of 0.5 points, with points allocated proportionally between the recommended level and the penalty point.

Table S2. Variables for the multiple imputation by chained equations

| Variable | Type of variable | Description of variable | Model used to predict missing data in this variable | N (%) with data on this variable |
| --- | --- | --- | --- | --- |
| **Outcome variables** |  |  |  |  |
| PyrMDS, age 46 years | Continuous | Range 0-15, with a higher score representing greater adherence to the Mediterranean diet pyramid | Linear regression | 5611 (45%) |
| **Exposure variable** |  |  |  |  |
| Early adulthood SETs, age 16-24 years | Categorical | Six classes of Continued Education, Managerial Employment, Skilled Non-manual Employment, Skilled Manual Employment, Partly Skilled Employment, and Economically Inactive | Not appliable | 12423 (100%) |
| **Mediators** |  |  |  |  |
| Household equivalent income, age 46 years | Continuous | The weekly household income divided by (1 + 0.5 x number of additional adults + 0.3 x number of children aged 0-15 years) | Linear regression | 7266 (58%) |
| Neighbourhood deprivation, age 46 years | Ordinal | Decile values of the Index of Multiple Deprivation Score 2015 | Ordered logistic  regression | 7923 (64%) |
| **Covariates** |  |  |  |  |
| Sex | Binary | Men and women | Not appliable | 12423 (100%) |
| PyrMDS, age 16 years | Continuous | Range 0-13, with a higher score representing greater adherence to the Mediterranean diet pyramid | Linear regression | 4079 (33%) |
| Exercise frequency, age 16 years | Ordinal | 0, 0-3, 3-5, >5 times/week of playing sports more than 30 minutes | Ordered logistic regression | 4571 (37%) |
| Sleeping hours, age 16 years | Continuous | Hours spent for sleeping in the previous day of the survey | Linear regression | 4437 (36%) |
| Smoking frequency, age 16 years | Ordinal | None, less than one, one to four, and 5 or more cigarettes/week | Ordered logistic regression | 4760 (38%) |
| Alcohol use, age 16 years | Continuous | Range 0-7, number of days having alcohol in the past week of the survey | Linear regression | 4284 (34%) |
| General health, age 16 years | Ordinal | Excellent, good, fair, and poor | Ordered logistic regression | 7052 (57%) |
| Malaise scale, age 16 years | Continuous | Range 0-22, with a higher score representing experiencing more symptoms associated with depression | Linear regression | 4760 (38%) |
| BMI, age 16 years | Continuous | Parental report of weight (kg) divided by the square of parental report of height (metres) | Linear regression | 4975 (40%) |
| Parental social class, age 10 years | Ordinal | Professional, managerial, skilled non-manual, skilled manual, partly skilled, and unskilled | Ordered logistic regression | 10344 (83%) |
| Parental education, age 10 years | Ordinal | No qualifications, trade apprenticeship/O-level, A-level, further vocational, and degree+ | Ordered logistic regression | 10490 (84%) |
| Household income, age 10 years | Ordinal | >250, 200-250, 150-200, 100-150, 50-100, 35-50, <35 pounds/week | Ordered logistic regression | 9824 (79%) |
| Family structure, age 10 years | Categorical | living with both parents, with one of the parents, and with others | Polytomous logistic regression | 10682 (86%) |
| Neighbourhood location, age 10 years | Categorical | Rural, country-in or close to village, outskirts or town or city, inner urban area, and council estate | Polytomous logistic regression | 10227 (82%) |
| Neighbourhood social rating, age 5 years | Ordinal | Range from 1 to 4, with a higher level representing a more well-to-do neighbourhood | Ordered logistic regression | 9826 (79%) |
| **Auxiliary variables for outcomes** |  |  |  |  |
| Days of having breakfast, age 42 years | Continuous | Range 0-7 | Linear regression | 7986 (64%) |
| Frequency of having convenience food, age 42 years | Ordinal | More than once a day, once a day, several times a week, once or twice a week, at least once a month, less often, and never | Ordered logistic regression | 8068 (65%) |
| Intake of fruit and vegetables, age 30 years | Ordinal | Servings of fruit and vegetables a day | Ordered logistic regression | 11153 (90%) |
| **Auxiliary variables for mediators** |  |  |  |  |
| NSSEC8, age 42 years | Ordinal | Range 1-8, from higher managerial, administrative and professional occupations to never worked and long-term unemployed | Ordered logistic regression | 8005 (64%) |
| Cohort member and partner’s take home income, age 42 years | Ordinal | Range 1-18, with a higher level representing higher take home income from all sources | Ordered logistic regression | 6277 (51%) |
| **Auxiliary variables for diet** |  |  |  |  |
| Intake of soft drink, age 10 years | Binary | Having soft drink every day, and less often | Binary logistic regression | 9890 (80%) |
| Intake of sweets, age 10 years | Ordinal | Nearly every day, quite often, sometimes, and hardly ever | Ordered logistic regression | 9890 (80%) |
| **Auxiliary variables for health covariates** |  |  |  |  |
| Frequency of playing sports, age 10 years | Ordinal | Never or hardly ever, sometimes, and often | Ordered logistic regression | 10623 (86%) |
| Sleep difficulty, age 10 years | Binary | Having sleep difficulty, and not having sleep difficulty | Binary logistic regression | 10654 (86%) |
| Smoking history, age 10 years | Binary | Having tried a cigarette, and never trying a cigarette | Binary logistic regression | 9828 (79%) |
| BMI, age 10 years | Continuous | Parental report of weight (kg) divided by the square of parental report of height (metres) | Linear regression | 9606 (77%) |
| Rutter behaviour scale, age 10 years | Ordinal | 0-80^th^, 81^st^-95^th^, and >95^th^ centile | Ordered logistic  regression | 10056 (81%) |
| Malaise Inventory score, age 26 years | Continuous | Range 0-24, with a higher score representing experiencing more symptoms associated with depression | Linear regression | 8355 (67%) |
| **Auxiliary variables for SEP covariates** |  |  |  |  |
| Father’s social class, birth | Ordinal | Range from 1 to 6: SC 1, SC 2, SC3 non-manual, SC 3 manual, SC4, SC5 | Ordered logistic regression | 10719 (86%) |
| Mother’s social class, birth | Ordinal | Range from 1 to 6: SC 1, SC 2, SC3 non-manual, SC 3 manual, SC4, SC5 | Ordered logistic regression | 10480 (84%) |
| Father’s age at completion of education,  birth | Continuous | Age in years | Linear regression | 11005 (89%) |
| Mother’s age at completion of education,  birth | Continuous | Age in years | Linear regression | 11398 (92%) |
| Family structure, age 5 years | Categorical | living with both parents, with one of the parents, and with others | Polytomous logistic regression | 10150 (82%) |

Note. To assist the imputation, we included the auxiliary variables as SEP at birth and at age 42 years, additional variables on diet and eating behaviours at ages 16, 30 and 42 years, and health variables at age 10 years.

Table S3. OLS regression results for the associations of early adulthood socioeconomic trajectories with adult diet quality at age 46 years, with sequential adjustment of covariates (n=12423)

|  | Estimated values of PyrMDS, β (95% CI) | | | |
| --- | --- | --- | --- | --- |
|  | Model 1 | Model 2 | Model 3 | Model 4 |
| *Early adulthood SET classes* |  |  |  |  |
| Continued education | Reference | Reference | Reference | Reference |
| Managerial | -0.57  (-0.69, -0.44) | -0.55  (-0.67, -0.43) | -0.48  (-0.61, -0.35) | -0.47  (-0.60, -0.34) |
| Skilled non- manual | -0.62  (-0.74, -0.50) | -0.69  (-0.80, -0.57) | -0.59  (-0.71, -0.47) | -0.58  (-0.70, -0.46) |
| Skilled manual | -0.91  (-1.05, -0.77) | -0.78  (-0.93, -0.64) | -0.66  (-0.82, -0.50) | -0.64  (-0.81, -0.48) |
| Partly skilled | -0.94  (-1.08, -0.79) | -0.84  (-0.99, -0.69) | -0.70  (-0.86, -0.54) | -0.70  (-0.87, -0.54) |
| Economically inactive | -0.86  (-1.04, -0.69) | -0.90  (-1.11, -0.70) | -0.74  (-0.96, -0.53) | -0.73  (-0.94, -0.51) |

Note. Model 1 is the unadjusted model, model 2 added the covariates of sex and adolescent health, model 3 further added the covariates of childhood SEP, and model 4, the fully adjusted model, added the covariate of adolescent diet quality.
